# Clinicians’ Perspectives on “Fighting the Ventilator”

**DOI:** 10.64898/2026.10.20.26365990

**Authors:** Lena Sophie Olivier, Denitsa Madzhakova, Camelia Lauterbach Oprea, André Stollenwerk, Thorsten Orlikowsky, Mark Schoberer

## Abstract

**Introduction:** “Fighting the ventilator” (FV) is characterized by a patient’s active counteraction of respiratory gas flow across multiple breaths during mechanical ventilation. An effective ventilation is prevented during such episodes. The objective of this exploratory study was to evaluate clinicians’ knowledge, perception and agreement regarding this condition.

**Methods:** In a cross-sectional online survey, medical professionals with experience in intensive care medicine (mostly neonatal and pediatric patients) responded to 11 questions covering various clinical aspects of FV.

**Results:** All respondents were familiar with the phenomenon and assessed it to be clinically relevant. Neonates were perceived to be predominantly affected. Promoting factors of FV included inadequate sedation and ventilation-associated factors. In clinical practice, the participants identified FV by clinical evaluation, analysis of ventilator waveforms and capnography and used both pharmacological and non-pharmacological intervention strategies. There was no consensus concerning the English terminology of the condition. Only few respondents were familiar with local guidelines to manage FV.

**Conclusion:** Although there was consensus on the relevance of FV, diagnostics and management strategies were highly heterogeneous. Many participants agreed that poor patient-ventilator synchrony was an important cause of FV. Future work should focus on characterizing and generating evidence on FV.

**Impact:**

- „Fighting the ventilator” is a commonly known and frequent condition in mechanically ventilated patients and is a sign of patient distress.
- This survey conducted among highly experienced clinicians (from NICUs and PICUs) highlights the substantial lack in standardized terminology and management strategies.
- 82% of participants considered the phenomenon highly relevant, but reported heterogeneous approaches concerning its recognition and therapeutic intervention.
- The majority were unaware of local or international guidelines to guide the management of “fighting the ventilator”.
- Future work should address these knowledge gaps and unify terminology. We advise against the use of the unscientific violence metaphor “fighting the ventilator”.

## Introduction

Although there is a shift from invasive to non-invasive ventilator support^1–3^, invasive mechanical ventilation remains a life-saving therapy in patients with respiratory failure, particularly in critically ill populations^4^. Adequate synchrony between the patient and the ventilator is essential for effective ventilation^5^. Patient–ventilator asynchronies (PVA) are a common complication of mechanical ventilation^6^ and may occur at different phases of the respiratory cycle, including triggering, flow delivery, and cycling^7,8^. PVA can lead to increased work of breathing, patient discomfort, prolonged duration of mechanical ventilation, and potentially worsen clinical outcomes due to inappropriate ventilation support^7,9,10^. Despite their high impact, they remain underdiagnosed^11,12^.

“Fighting the ventilator” (FV) represents a distinct and clinically challenging complication of mechanical ventilation. It describes a condition in which a ventilated patient counteracts the respiratory gas flow applied by the ventilator across multiple consecutive breaths, thereby impairing effective ventilation^13^. This may result in reduced tidal volumes and impaired gas exchange^14^ and can both be cause and result of patient distress. Despite its potential clinical relevance, recognition of FV in routine care remains challenging, partly due to inconsistent terminology and the absence of standardized diagnostic criteria^15,16^. This may contribute to a variability in bedside recognition and management. Furthermore, little is known about how clinicians perceive and identify FV in routine care.

We were motivated to conduct this study because we had the impression that practically every caregiver on the intensive care unit (ICU) is familiar with this problem and regards it as relevant in personal communications with colleagues. At the same time we found that this particular form of PVA is distinct from those which are established in literature. Only little literature can be found; there is no appropriate and uniform medical terminology, and it is not included into medical guidelines. To substantiate these assumptions, we performed an interdisciplinary exploratory survey among experienced caregivers, mostly from neonatal and pediatric ICUs.

Specifically, we sought to assess the terminology used in order to include it into a scoping review. We furthermore evaluated perceived risk factors, clinical signs, diagnostic strategies, and therapeutic interventions associated with FV.

Within this manuscript, we use the term “fighting the ventilator” despite our concerns regarding its adequacy. These are substantiated in the discussion section. The rationale for its use is our belief that there is currently no term in broader use for this condition among intensive care specialists, and because it differentiates the condition from other PVAs.

In the German-speaking community, the term “Pressen” (pressing) is most frequently used, and was therefore also used for this survey in its original form. Pressing refers to the muscular activation which is often perceived to be the “fighting” element in FV.

## Methods

### Study design

The study was conducted as an explorative cross-sectional online survey. The target population comprised German-speaking medical professionals of different occupational groups experienced in neonatology and/or pediatric/adult intensive care.

### Questionnaire development and pre-testing

The questionnaire was developed by three authors with the aim to investigate clinically relevant aspects of FV, as well as its perception among medical professionals. The survey consisted of twelve questions from ten different thematic areas. All questions were phrased in German language. One question addressed German terminology and was excluded from the present paper. The questionnaire was structured into four sections, with two to four questions being presented per page. Both free-text options and single or multiple-choice answers were provided. An English translation of the questionnaire is presented in Table 1.

**Table 1:**
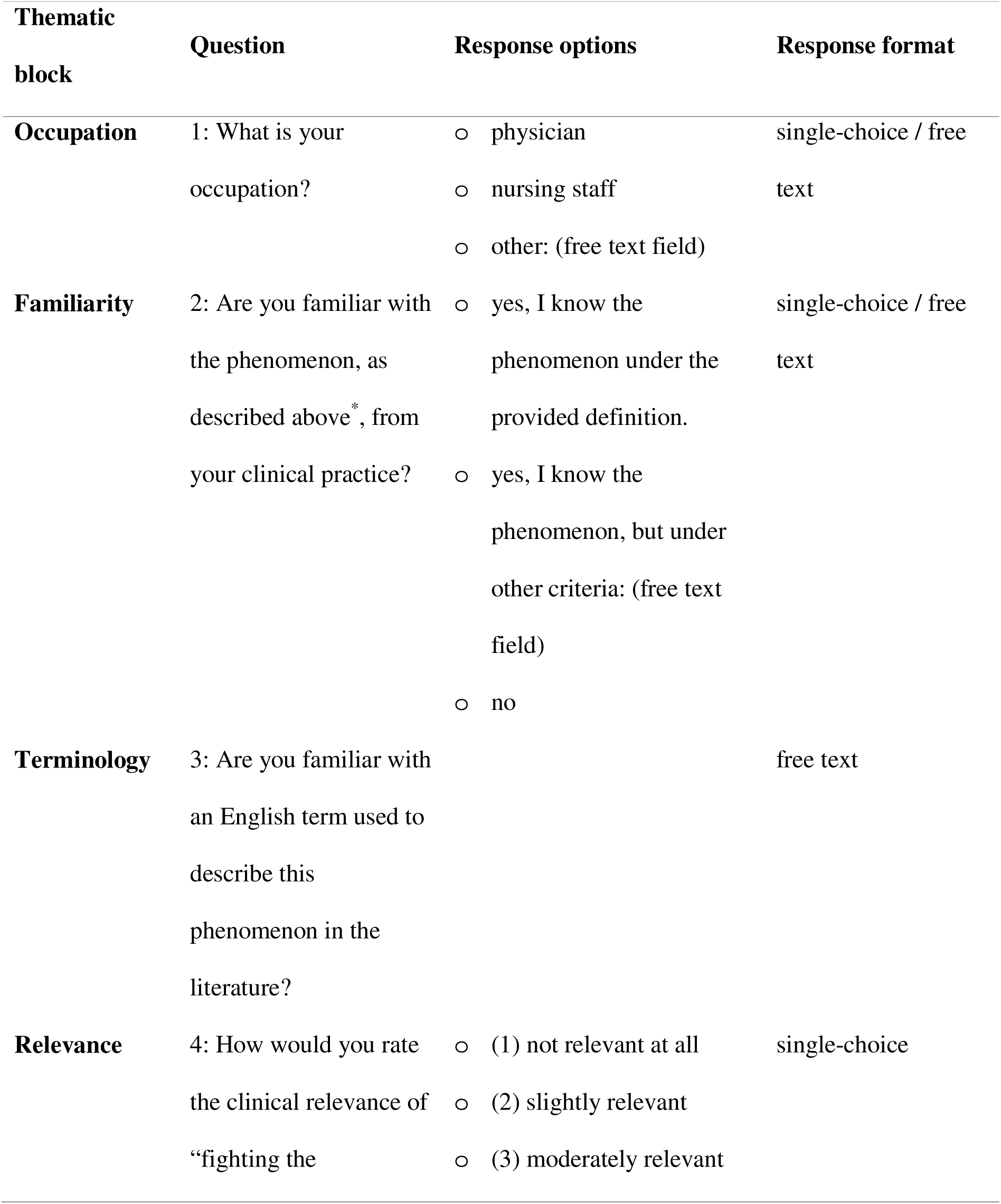
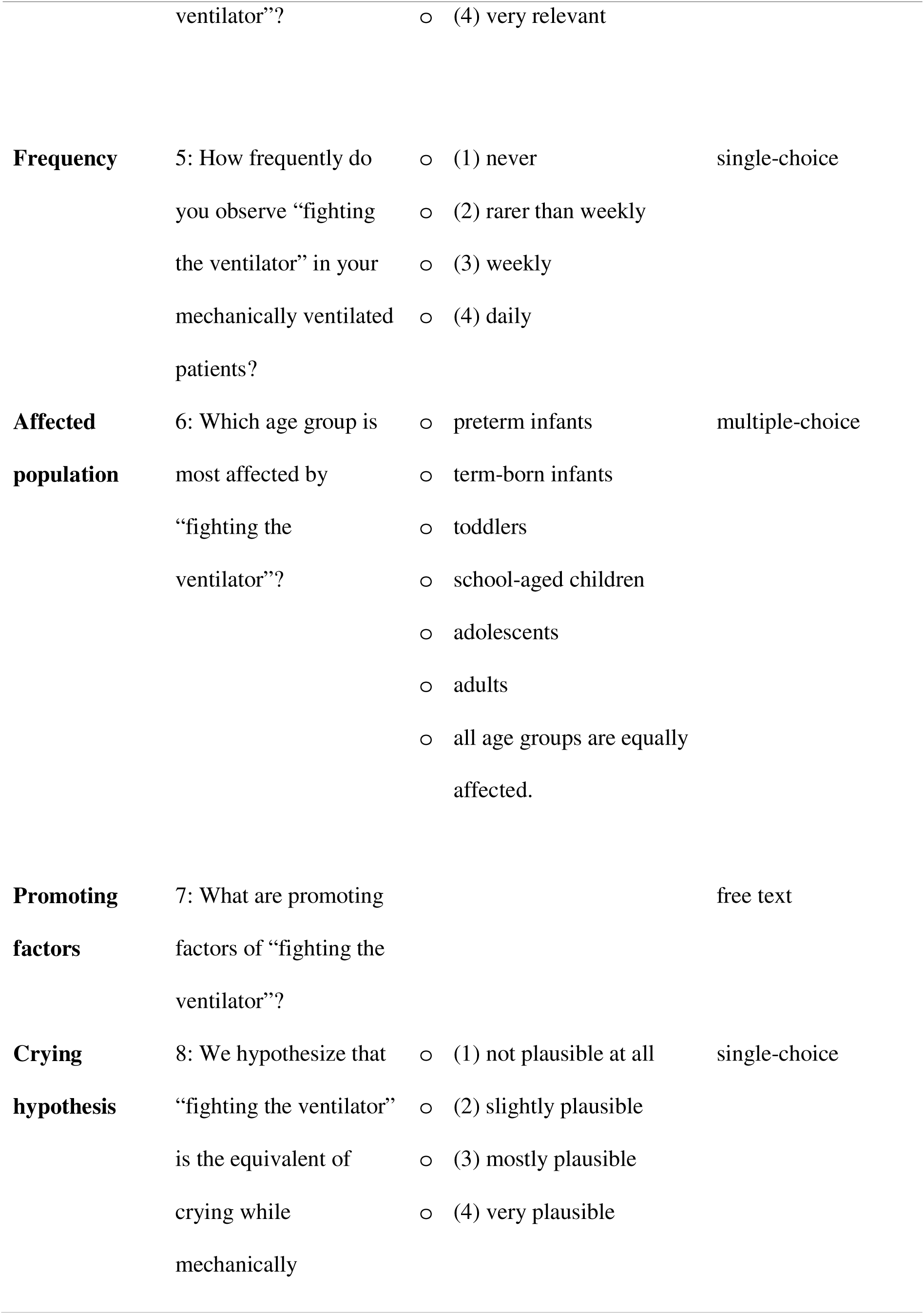
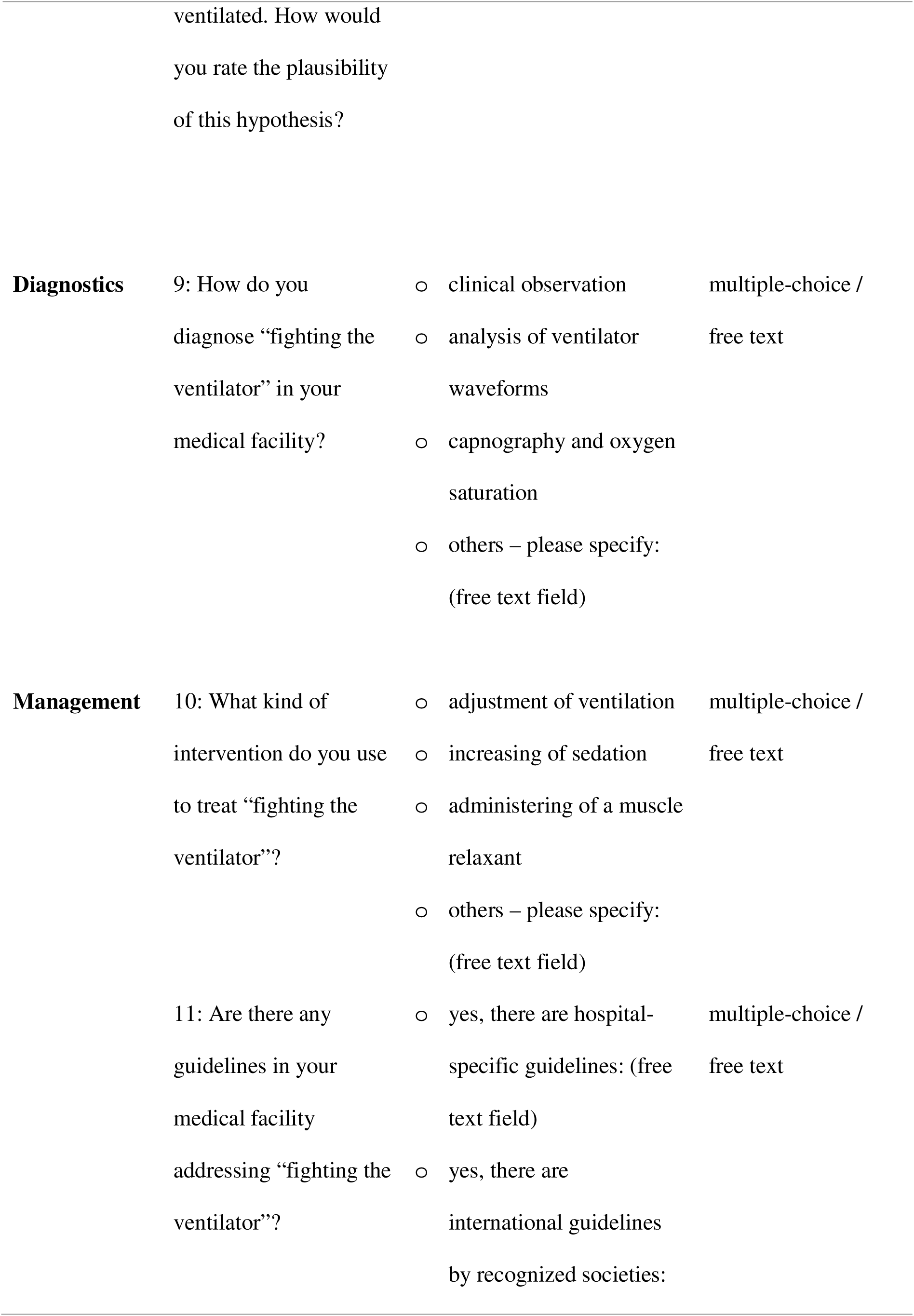
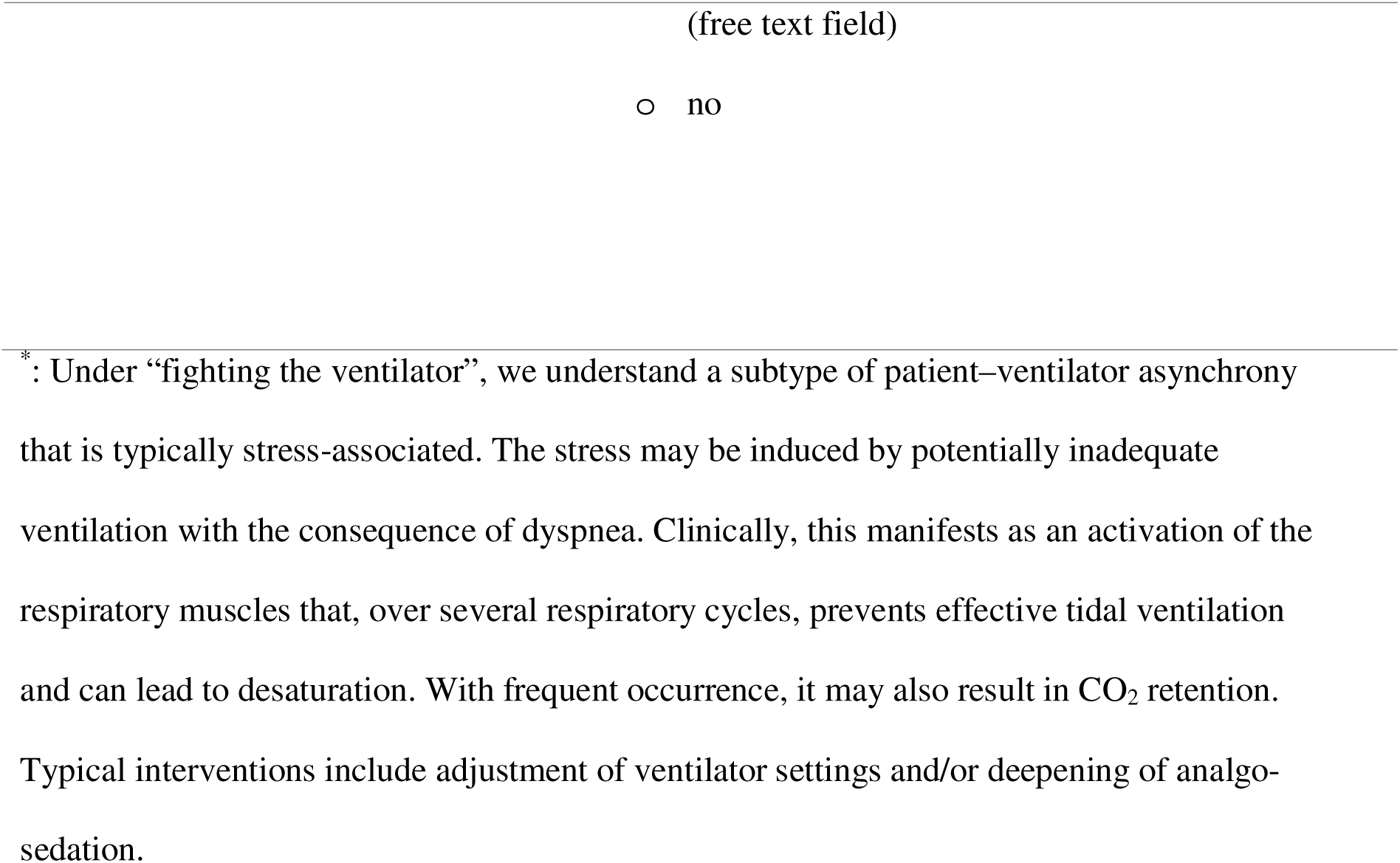
Questionnaire on “fighting the ventilator”. The response options of ordinal items (relevance, frequency and crying hypothesis) were assigned ranks from 1 to 4 with increasing strength of agreement.

To uncover implausibility and functional problems, two pre-tests were iteratively conducted, each independently by two neonatologists from the team.

### Recruitment process

The study was conducted as a closed survey. The participants were contacted via institutional e-mail addresses, containing a personalized (single-use) link to the survey. This approach ensured that only medical professionals with the required qualifications responded to the survey. A reminder e-mail was sent to participants who had not yet started or completed the survey within two weeks of the invitation.

### Survey administration

The web-based software SoSci Survey (SoSci Survey GmbH, Munich, Germany) was used to create the online questionnaire and collect the responses. The survey was accessible from 6 February 2026 to 31 March 2026. The participation was voluntary with no incentives being offered.

To obtain informed consent, respondents were provided with detailed information about the study, including its objectives, data protection practices and estimated completion time, prior to participating in the survey. A suggested definition of FV was provided to assess whether the participants were familiar with the phenomenon (see Table 1, footnotes). Adaptive questioning was used to ensure that respondents not familiar with FV (question 2, see Table 1) did not have to respond to subsequent questions related to the topic. With exception of the first question, none of the items required obligatory answering. The respondents were able to review and change their answers using the webpage’s back button before completing the questionnaire.

### Data handling and analysis

All data were collected, processed and analyzed in a pseudonymized fashion. No personal information was collected. Participants were not re-contacted. Empty responses were treated as missing values and excluded from the analysis. The data preparation was performed using Microsoft Excel (Microsoft Corporation, Redmond, WA, USA). Responses were coded on a four-point ordinal scale ranging from 1 (lowest ranking) to 4 (highest ranking) where appropriate (questions regarding relevance, frequency and crying hypothesis). Data analysis was conducted using IBM SPSS Statistics, version 31.0.0.0 (IBM Corp., Armonk, NY, USA). Free-text responses were grouped into content-based categories using an inductive coding approach by one author and checked by another^17^. Descriptive analyses were carried out to summarize the data. Group comparisons were carried out between physicians and nursing staff. Differences between these groups were assessed using the Mann-Whitney-U-test and Fisher’s exact test where appropriate. Statistical significance was set at a two-tailed 0.05-level.

### Ethical approval

The survey was approved by the local ethics committee (Ethik-Kommission an der Medizinischen Fakultät der Rheinisch-Westfälischen Technischen Hochschule Aachen (RWTH Aachen), EK 25-199). The overall project AIx-Neo-Guard^18^ was registered prospectively at the German Clinical Trial Register (DRKS00033228). The survey was conducted in accordance with the declaration of Helsinki^19^.

## Results

### Respondent characteristics

Of 66 contacted individuals, 44 completed the questionnaire (response rate 66.7%). The mean completion time was 5 minutes 30 seconds. 20 respondents (45.5%) were female. 36 participants (81.8%) were physicians and 8 (18.2%) were nursing staff members. Professional roles among physicians comprised clinical directors (n=4; 11.1%), senior physicians (n=23; 63.9%) and board-certified specialists (n=9; 25%). The physicians’ disciplines are listed in Table 2, with most physicians being specialized in more than one discipline. The data were obtained from 21 hospitals across 20 cities. Most participating hospitals were located in Germany (n=18), followed by the Netherlands (n=2) and Switzerland (n=1). 14 institutions were university hospitals.

**Table 2:** Specialties among participating physicians (multiple specializations possible)

| **Specialty** | **Respondents, *n* (%)** |
| --- | --- |
| Neonatologists | 27 (75.0%) |
| Pediatric intensive care physicians | 21 (58.3%) |
| Pediatric cardiologists | 2 (5.6%) |
| ICU–specialized anesthesiologists | 6 (16.7%) |

### Familiarity with FV and relevance

43 participants (97.7%) were familiar with the phenomenon under the provided definition in the questionnaire (Table 1, see footnotes). One respondent was familiar with FV but provided an alternative definition, describing the insufficient ventilation as a consequence rather than the cause of FV.

The relevance of FV was deemed moderate (n=8; 18.2%) to high (n=36; 81.8%) among the participants, without a statistically significant difference between physicians and nursing staff (Mann-Whitney U = 134.00, p = 0.78).

### English terminology

The distribution of responses regarding English terminology is illustrated in Figure 1. Half of the participants were unaware of an established English term for FV. The most frequently reported term was “patient-ventilator asynchrony” (n=10; 22.7%), followed by “fighting the ventilator” (n=8; 18.2%).

**Figure 1:**
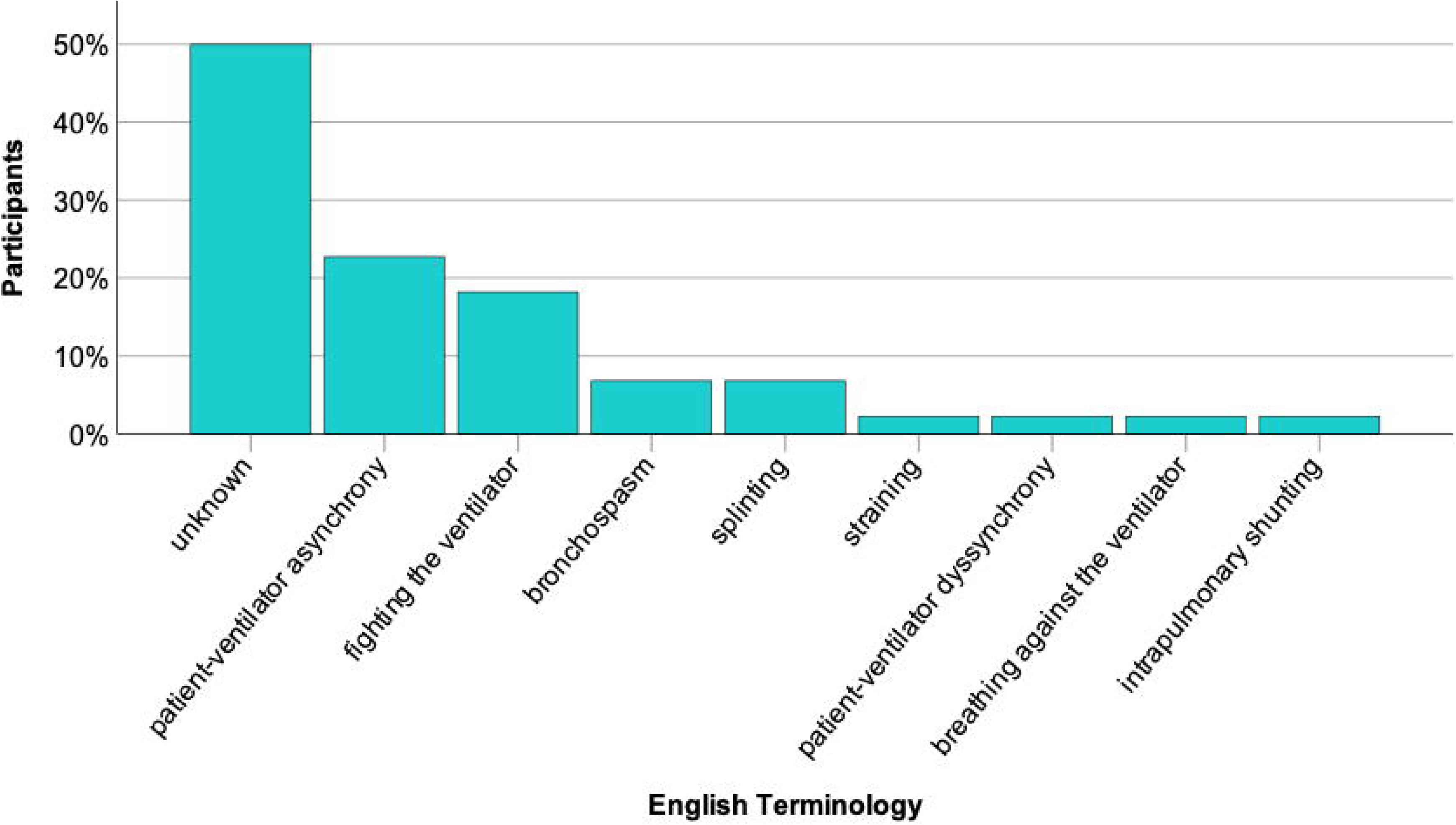
Distribution of reported English terminology of FV

### Frequency

Overall, 86.4% of respondents (n=38) reported observing the phenomenon at least once per week. Of these, 45.5% reported weekly observation of FV, while 40.9% observed FV daily. The remaining participants (n=6; 13.6%) estimated the occurrence of FV as less frequent than weekly. None of the respondents selected the response option “never”. The perceived frequency of FV did not differ significantly between physicians and nursing staff (Mann-Whitney U = 140.00, p = 0.92).

### Affected Population

Premature infants were most frequently identified as being affected by FV (n=26; 59.1%), followed by term-born infants (n=23; 52.3%). The response frequencies for the age groups are presented in Table 3.

**Table 3:**
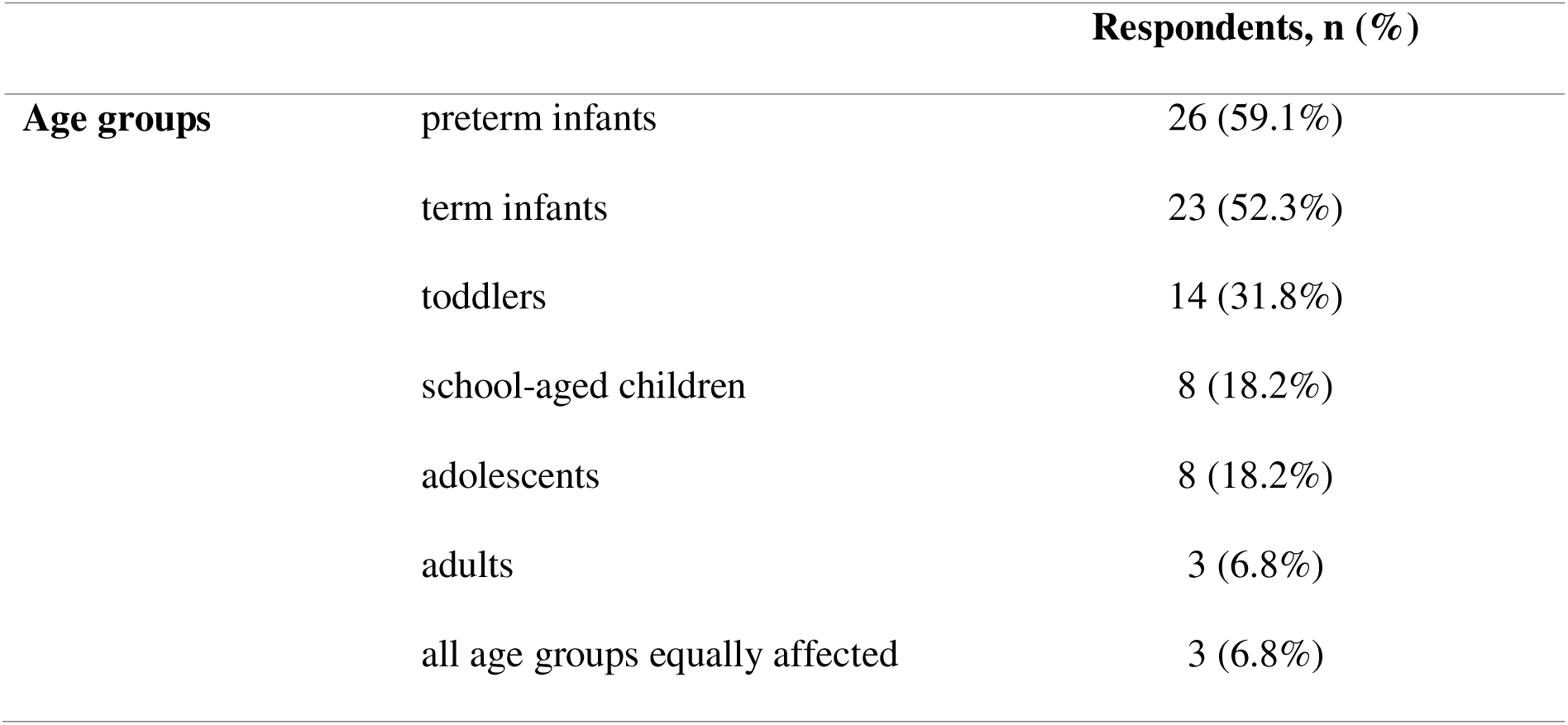
Distribution of perceived affected age groups (multiple selections possible)

### Promoting factors

Free text responses were grouped into categories. The most frequently reported categories for promoting factors were “insufficient sedation” (52.3%), “ventilation-associated factors” (45.5%) and “respiratory/pulmonary impairment” (31.8%). Further associated factors comprised pain, external stimuli and stress. An overview of the responses is given in Figure 2.

**Figure 2:**
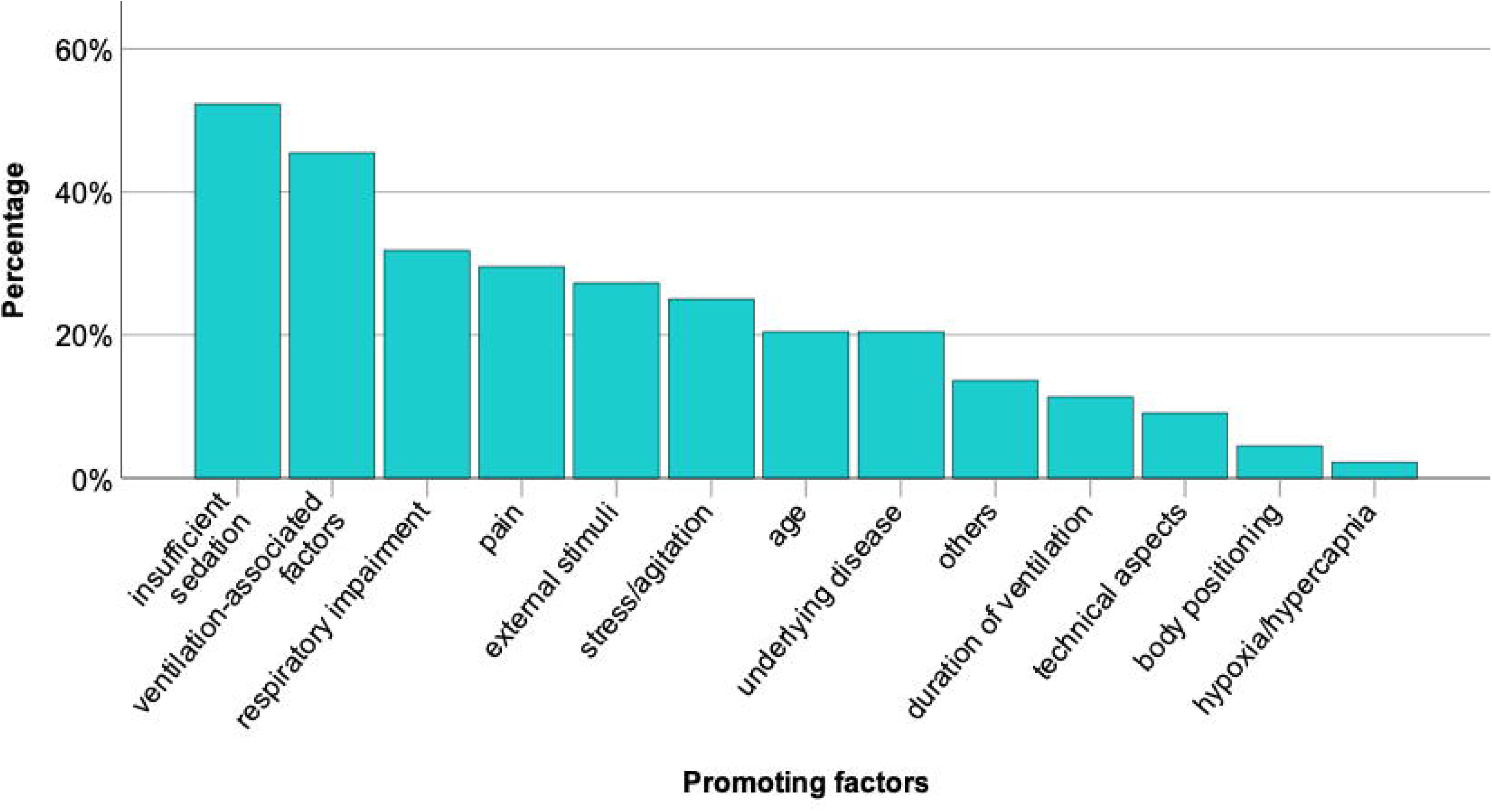
Most frequently reported promoting factors of FV, shown as percentage of entries for each category. The category “others” comprised the following responses: breathing against the ventilator, recovery from anesthetic and “on a whim”

### Crying hypothesis

The crying hypothesis was deemed very or mostly plausible by 25 participants (n=25; 56.8%). 18 participants (40.9%) considered the hypothesis to be slightly plausible, while one respondent (2.3%) rated it as not plausible at all. No statistical difference was observed between the occupational groups (Mann–Whitney U = 139.00, p = 0.89).

### Diagnostics

The participants were asked how they diagnose FV in their institution, with multiple responses permitted. All participants reported using clinical observation to detect FV. Ventilator waveform analysis was selected by 24 respondents (54.6%), while 21 respondents (47.7%) reported using capnography and/or oxygen saturation monitoring. 6.8% of responses (n=3) were categorized as “others”, including “decrease of systolic blood pressure, increase in central venous pressure”, “heart rate and blood pressure monitoring” and “relation between tidal volume and pressure during mechanical ventilation”. No statistically significant differences were observed between the occupational groups for any of the diagnostic tools proposed in the questionnaire, including ventilator waveform analysis (p=1.00; Cramér’s V=0.04), capnography/oxygen saturation monitoring (p=0.45; Cramér’s V=0.14) and other diagnostic approaches (p=0.46; Cramér’s V=0.11).

### Management

The most frequently reported management strategies included adjustment of ventilation (n=40; 80.9%) and the increase of sedation (n=40; 90.9%). 13 participants (29.5%) reported to use muscle relaxants to alleviate FV. Free-text entries were provided by 15 respondents (34.1%). The most common free-text responses were suctioning (n=5), extubation (n=4), soothing and comfort (n=4) as well as additional diagnostic procedures (n=2). 3 responses which did not correspond to the mentioned categories were: “circulatory stabilization (inotropes, fluids, calcium)”, “pain management” and “positioning”.

We categorized the management strategies as pharmacological (“increase of sedation” and “muscle relaxation”) or non-pharmacological (“adjustment of ventilation”). Most participants (n=36; 81.8%) reported using both pharmacological and non-pharmacological management approaches. The remaining participants (n=8; 18.2%) reported using only one approach, with 4 participants (9.1%) selecting exclusively pharmacological approaches and 4 participants (9.1%) selecting exclusively non-pharmacological management strategies. Overall, both pharmacological and non-pharmacological strategies were reported by 40 participants respectively (90.9%).

### Guidelines

Only 4 respondents (9.1%) reported using local standard operating procedures to guide the management of FV. These were specified in the free text responses as “guideline for clinical observation and extubation criteria”, “guideline for ventilation”, “guideline for ventilation and sedation”, and “apnea of prematurity guideline”. The remaining participants (n=40; 90.9%) reported being unaware of local or international guidelines for the management of FV.

## Discussion

### Key results

All respondents were familiar with FV and considered the phenomenon to be clinically relevant. FV was assessed to be a frequent condition. Respondents perceived that especially neonates are affected and reported a variety of possible risk factors, such as inadequate sedation or ventilator-associated factors. FV detection was based on clinical evaluation, the analysis of ventilator waveforms, pulse oximetry and capnography. The most frequently reported interventions were pharmacological treatment (sedation, relaxation), adaption of ventilator settings and reduction of stressors. Less than 10% of respondents used local guidelines for FV management. No internationally recognized guidelines were identified. Knowledge about English terminology was, limited and heterogeneous.

### Interpretation

PVA is an umbrella term which typically encompasses seven distinct entities: ineffective efforts during expiration, double triggering, premature cycling, delayed cycling, reverse triggering, flow starvation and auto-cycling^20^. FV represents a specific form of abnormal patient–ventilator interaction. It is characterized by the impairment of effective mechanical ventilation across multiple breaths. The patient counteracts the respiratory gas flow applied by the ventilator. To date, FV (or any other term describing a patient-generated cross-cycle PVA) is not mentioned in the established list of PVA types.

All survey participants perceived FV as a clinically relevant condition, yet there was considerable disagreement regarding English terminology: Eight different terms were suggested. 50.0% of participants were unaware of an established English term. 25.0% of respondents used “patient-ventilator asynchrony / dyssynchrony” and 18.2% “fighting the ventilator”, which are both suitable umbrella terms but do not specifically describe the pathophysiology of FV. This underlines the lack of consensus regarding terminology of FV and the heterogeneity of used terms.

Kim et al. previously advised against the use of the term “fighting the ventilator” because violence metaphors related to patients perpetuate negative stereotypes and stigmata^21^. We agree to this view and believe that the unscientific and non-medical terminology might even impede a systematic, scientific occupation with and the diagnostic perception of the phenomenon. Instead, part of the literature refers to the phenomenon as “hypoxemia episodes” or “desaturation” ^16,22^. These terms, despite being much more adequate in our opinion, lack in specificity and describe the consequences rather than the underlying mechanism. Hypoxemia and desaturation can originate from various conditions, e.g. apnea of prematurity^23^ or pneumonia^24^.

We propose the use of the descriptive term “cross-cycle impairment of mechanical ventilation”, which arose from discussions within our research team. We believe that it constitutes a specific description of the phenomenon, which still leaves room for further research on the underlying mechanisms (e.g. reflectory, muscular or intentional). However, we only suggest this term with the aim to open a discussion within the research community. International consensus should harmonize terminology to facilitate comparison across studies and to allow to diagnose the condition.

Our survey found that 45.5% of respondents agreed that inadequate ventilator settings can promote FV. This suggests that poor patient–ventilator synchrony may be an important cause of FV, analogous to how ventilator settings contribute to the established forms of PVA^25^. PVA are predominantly diagnosed by ventilator waveform analyses^7,26^. Interestingly, our results indicate that diagnostic strategies for FV are more heterogeneous. While 54.6% of respondents also employed ventilator waveform analysis for FV detection, clinical observation was used by all respondents, and half of the participants additionally used capnography. Former studies demonstrated that the detection of established forms of PVA can be improved with targeted training^12,27^. As FV is primarily detected through clinical observation, there is a need for specific educational materials on recognizing and managing FV, including waveform signatures, capnography patterns and bedside signs.

Multiple studies addressing established PVA agree that PVA are common and associated with serious complications (e.g. prolonged mechanical ventilation, higher rates of tracheostomy, mortality)^20^. In our study, the clinical impact of FV was also assessed to be high. However, the long-term effects of FV remain unknown and insufficiently characterized. We hypothesize that both short- and long-term consequences of FV are clinically important and require further investigation.

### Limitations

The objective of this study was to conduct an exploratory survey on clinicians’ perspectives on FV. We therefore used a small and non-representative convenience sample. However, the average clinical and scientific experience of the respondents was high.

Nurses and medical personnel treating adult patients were underrepresented. The small sample size did often not allow for a robust comparison of the subgroups (e.g. physicians vs. non-physicians). While we received responses from multiple institutions and three different European countries, the international perception of FV remains uncharacterized. Future studies should also explore transnational differences using larger sample sizes, which will help to establish international guidelines.

The questionnaire partially provided multiple choice options. Predefined response options may have prompted responses that would not have emerged in free-text answers (recognition bias).

## Conclusion/Outlook

Respondents broadly agreed that FV is a relevant and frequent phenomenon in mechanically ventilated patients. Multiple risk factors were identified. Diagnostics and management strategies were highly heterogeneous, indicating that the phenomenon is insufficiently characterized. Future work should focus on establishing a consistent terminology and on generating more evidence to allow the development of a more standardized therapeutic approach and ideally international guidelines.

## Declarations

## Data availability statement

The datasets used and analyzed during the current study are available from the corresponding author on request.

## Acknowledgements

The authors wish to thank all participants of the survey for their thorough responses and valuable input. We deliberately chose not to name the participants in this context to respect data protection regulations. The authors furthermore wish to thank Sarah Essers, Milian Brasche and Jacob Eschweiler for their assistance in the participant recruitment process and Jan Wienströer for technical assistance with the survey.

## Consent statement

Informed consent was obtained from the participants prior to responding to the survey.

## Ethics approval

The project was approved by the local ethics committee (Ethik-Kommission an der Medizinischen Fakultät der Rheinisch-Westfälischen Technischen Hochschule Aachen (RWTH Aachen), EK 25-199).

## Consent for publication

not applicable

## Author contributions

**LO**: Conceptualization, Data curation, Formal Analysis, Investigation, Methodology, Resources, Supervision, Validation, Visualization, Writing – original draft, Writing – review & editing;

**DM**: Conceptualization, Data curation, Formal Analysis, Investigation, Methodology, Writing – original draft;

**CLO**: Software, Validation, Writing – review & editing;

**AS**: Funding acquisition, Project administration, Software, Validation, Writing – review & editing;

**TO**: Writing – review & editing;

**MS**: Conceptualization, Data curation, Funding acquisition, Investigation, Methodology, Project administration, Resources, Supervision, Validation, Writing – review & editing

